# Social contacts and other risk factors for respiratory infections among internally displaced people in Somaliland

**DOI:** 10.1101/2021.08.20.21262338

**Authors:** Kevin van Zandvoort, Mohamed Omer Bobe, Abdirahman Ibrahim Hassan, Mohamed Ismail Abdi, Mohammed Saed Ahmed, Mohamed Yusuf Warsame, Muna Awil Wais, Emma Diggle, Catherine Satzke, Kim Mulholland, Mohamed Mohamoud Egeh, Mukhtar Muhumed Hassan, Mohamed Abdi Hergeeye, Rosalind M Eggo, Francesco Checchi, Stefan Flasche

## Abstract

**Background:** Populations affected by humanitarian crises experience high burdens of acute respiratory infections (ARI), potentially driven by risk factors for severe disease such as poor nutrition and underlying conditions, and risk factors that may increase transmission such as overcrowding and the possibility of high social mixing. However, little is known about social mixing patterns in these populations.

**Methods:** We conducted a cross-sectional social contact survey among internally displaced people (IDP) living in Digaale, a permanent IDP camp in Somaliland. We included questions on household demographics, shelter quality, crowding, travel frequency, health status, and recent diagnosis of pneumonia, and assessed anthropometric status in children. We calculated age-standardised social contact matrices to assess population mixing, and conducted regression analysis on risk factors for recent self-reported pneumonia.

**Results:** We found crowded households with high proportions of recent self-reported pneumonia (46% in children). 20% of children younger than five are stunted, and crude death rates are high in all age groups. ARI risk factors are common, but we did not find any significant associations with self-reported pneumonia. Participants reported around 10 direct contacts per day. Social contact patterns are assortative by age, and physical contact rates are very high (78%).

**Conclusions:** ARI risk factors are very common in this population, while the large degree of contacts that involve physical touch could further increase transmission. Such IDP settings potentially present a perfect storm of risk factors for ARIs and their transmission, and innovative approaches to address such risks are urgently needed.

## Introduction

Prior to the COVID-19 pandemic, acute respiratory infections contributed to over 2 million deaths annually.^1^ These pathogens are transmitted via direct or indirect (e.g. via fomites) contact with respiratory droplets. Unsurprisingly, patterns of transmission for respiratory pathogens correlate strongly with social contact patterns among different groups of people.^2,3^ In addition, other factors such as malnutrition^4^ modulate host responses to infection thereby affecting both individual susceptibility to infection and the rate of pathogen shedding, i.e. infectiousness to others.

Accordingly, epidemiological transmission models commonly stratify populations into groups, usually by age, and assume different contact rates among these groups. To accurately parametrize such models, several surveys have collected empirical contact data by asking participants to report the frequency with which they come into direct contact with people from their and other groups.^5–9^ The majority of these studies have been conducted in high-income countries, with limited information from lower- and middle-income countries.^10^ To our knowledge, no contact surveys have captured the experience of populations living in refugee or internally-displaced persons’ (IDP) camps, despite these settings experiencing a high burden of endemic and epidemic-prone infections, including respiratory diseases.^11,12^

Nearly 80 million people were forcibly displaced due to insecurity and war in 2019.^13^ Many of these people live in overcrowded camps where residents are often unemployed or unable to work, not all children may have access to school, hygiene may be curtailed by inadequate water and sanitation services, and people may be required to gather in large groups during relief distributions or at queues at water points, etc. These factors likely affect social contact patterns, in turn impacting the spread of infections. Food insecurity and resultant acute malnutrition are also a major threat, particularly among IDPs, and increase both transmission and case-fatality of most respiratory infections.^14^

Epidemiological transmission models are an increasingly appreciated tool to quantify infection transmission and efficiently explore the impact of possible interventions. In the context of a project to study the effectiveness of pneumococcal conjugate (PCV) vaccination among IDPs, we aimed to parameterise a model of *Streptococcus pneumoniae* transmission.^15^ We therefore conducted a contact and risk factor survey in an IDP camp in Somaliland in 2019 to quantify factors commonly associated with transmission of respiratory infections.

## Methods

### Study population

We conducted a cross-sectional survey on social contacts and prevalence of risk factors in the Digaale IDP camp near Hargeisa, the capital of Somaliland. The camp was established in 2014, in response to a large influx of displaced people in Somaliland following an acute food insecurity crisis. It is situated 3km from Hargeisa airport and 4km from the city borders, has an area of 15 hectares and is surrounded mainly by desert and shrubland. Digaale has a small primary healthcare centre, that operates under the Somaliland Ministry of Health Development, and a primary school. There are 894 shelters in Digaale that are all constructed from corrugated sheets. Each household has access to a private latrine and private water tank. Several photos of Digaale IDP camp are provided in section A of the supplementary material.

### Study design and sample selection

The survey was nested within a larger study of pneumococcal carriage, and aimed to include a representative sample of 100 individuals in each of the following age groups: <1, 1, 2-5, 6-14, 15-29, 30-49, 50+ years old. Purposes and procedures of the study were presented to and discussed with the elders and other representatives from the community in advance. Data collection was conducted by the Somaliland Ministry of Health Development in collaboration with Save the Children International, who provide health and nutrition services in the camp.

A team of 12 enumerators were trained for a week and undertook data collection between October-November 2019. Data collection was done from 8AM until 4PM on all days of the week except Friday. A community leader notified the population living in the block that would be visited on specific days, and answered any questions regarding the presence of enumerators. Adults provided informed written consent, including on behalf of their children under the age of 18, and acted as proxies for young children who were unable to answer for themselves. Children aged >12 and <18 years were asked for assent.

As there was no sampling frame, we employed quota sampling to reach the desired sample size in each age group. We visited all shelters in Digaale and conducted a household survey in consenting households. A household was defined as individuals who live together and share a common source of food. The survey followed the Standardised Monitoring and Assessment of Relief and Transitions (SMART) guidance to collect the age and gender of those currently living in the household, and retrospectively assessed who had been born or died, and who migrated in or out of the household in the preceding six months.^16^ A localized event calendar was used to aid in the recollection of timing of events. In addition, the survey included questions to ascertain the presence of household level risk factors: total number of rooms, leakage or draft in the shelter, cooking fuel used, ventilation used in the cooking area, water source, and substance use in the household. At completion of the survey, household members were assigned sampling weights based on their age. These weights were then used to sample one household member, who was invited to take part in the second stage of the survey. We asked those consenting to remember all people they would contact on the following day, and agreed a time for a return visit on the day thereafter.

During the return visit, we conducted a second survey that assessed individual travel patterns and direct social and physical contacts. The survey was developed as an extension of contact surveys conducted in non-IDP populations.^5,6^ Participants were asked to first list (nick)names or initials of all their contactees. A contactee was defined as any individual who was met in person during the 24 hours before waking up on the day being surveyed, and with whom the participant had at least a short conversation (direct contact). For each contactee, participants were asked to list several characteristics: estimated age (in years) and gender, relationship to the contactee, setting where the contact occurred, type of contact (physical touch or nonphysical), duration of the contact, and typical frequency with which participant and contactee have contact. Physical touch was defined as any form of skin-to-skin contact. Participants were also asked to estimate the total number of indirect in-person contacts they made that did not fit the criteria for a direct contact, and to list any health conditions diagnosed by a health professional. If pneumonia was listed as a health condition, we asked whether diagnosis occurred within the six months preceding the survey.

For all participants aged 6-59 months, we measured height (or length if less than 85cm), weight, middle-upper arm circumference (MUAC) and presence of bilateral oedema on the dorsum of both feet as described in the SMART guidance. Anthropometric instruments and observations were standardised prior to data collection on a convenience sample of children seen at the health facility.

Shelters where no individual was present on the first visit were revisited on different days throughout the study period, up to a total of five times. To assess selection bias for shelters where no person was present on all visits, we asked neighbours of a random sample of 96 of these shelters whether their neighbouring shelter was still inhabited.

Participants who were not available on the return visit were also revisited up to a total of five times. If participants were no longer available, or withdrew consent, they were replaced with another individual of the same household within the same age group, where possible.

All data were collected on electronic tablets using Open Data Kit software^17^. Ethical approval for the study was granted by the Somaliland Ministry of Health Development, Directorate of Planning, Policy, and Strategic Information, and by the London School of Hygiene & Tropical Medicine.

### Data analysis

As the sample for the participants included in the second phase of our survey was not self-weighting, and we deliberately oversampled individuals in the youngest and oldest age groups, we used post stratification weights to calculate population representative estimates of individual-level data.^18^ These weights are the inverse of a participant’s probability of selection in the sample, which was the product of the inverse of their household size and the estimated proportion of the population included in the sample in their respective age- and gender stratum. We further applied an additional weight to correct for imbalances in the distribution of days of the week within the final survey sample when estimating contact rates. We censored sample weights below or above the 5th and 95th percentile of all sample weights to those percentiles.

As we sampled a large proportion of households (65%) and individuals (17%) living in Digaale, we used finite population corrections (FPC) to calculate standard errors. The total population size of Digaale used in the FPC, for all ages and within each age group, was estimated by multiplying the total number of household members reported in the survey with a correction factor. This factor was calculated as the survey-estimated proportion of inhabited shelters included in the survey, the latter being the upper bound of the survey-estimated proportion of non-abandoned shelters. More detailed information is provided in Supplemental Section B. The *survey* package in R was used to perform the weighting and to apply the FPC when estimating weighted means, proportions, and quantiles where applicable.^19^

To estimate birth, death, and migration rates, we calculated person-time for each reported household member, including those who had died or migrated out, by assigning six months (the full recall period) to any individual who lived continuously within the household over the period. As we did not record exact dates when deaths, births and migration events occurred, an individual’s person-time was reduced by 50% for each of these events experienced. We estimated weighted rates by fitting a Poisson generalized linear model using the natural logarithm of person-time as an offset.

Nutritional data were analysed by calculating age- and sex-standardised z-scores for a range of anthropometric indices based on the World Health Organization Growth Reference standards^20^, which were subsequently categorized to assess malnutrition status. The *zscorer* package in R was used for the anthropometric analyses.^21^

We constructed contact matrices to visualise age-stratified daily contact rates and per capita contact rates, which were adjusted for reciprocity in the total number of contacts using the method by Wallinga et al.^22^ Uncertainty in the contact matrix was quantified by taking 10,000 bootstrap samples of all participants in the survey.

The relationship between different risk-factors and the cumulative incidence of self-reported pneumonia was assessed using unweighted logistic regression, adjusted for age and sex as a-priori defined confounders. We did not apply an FPC in computing the standard errors of the regression estimates.

All analyses were conducted in R 4.0^23^. Analysis scripts, anonymized data, and questionnaire scripts are available on GitHub via https://github.com/kevinvzandvoort/espicc_somaliland_digaale_contact_survey_2019. The anonymized contact data has also been uploaded to the respective repository on Zenodo (https://zenodo.org/communities/social_contact_data) to be accessible through the *socialmixr* package in *R* for epidemiological contact surveys.^24^

## Results

We visited all 894 shelters in Digaale IDP camp. Of these, 405 were empty on all occasions visited. We randomly sampled 96 of these empty shelters and asked neighbours whether the shelter was occupied. Using this sample, we estimate that 50% (95%CI 39-60) of the shelters had been uninhabited for a long time, while 38% (28-49) of the shelters were occupied and 12% (6-19) were a shop or combined with another shelter already included in the survey. Using the upper bound of these estimates, we conservatively assume that there are a total of 872 unique shelters in Digaale, of which 715 are inhabited.

Twenty-five households declined consent. We thus enrolled and collected demographic information from 2,049 individuals who were living in 464 households at the time of the survey (65% of inhabited unique shelters), with additional information collected about 166 individuals who migrated out of these households and 34 individuals who had died in the six months preceding the survey. In the contact survey we enrolled 509 participants from 426 households, who provided information about 4,857 contactees. Of all participants included in the contact survey, we collected anthropometric estimates from 171 children aged 6 to 59 months. Despite their inclusion and consent in the first household visit, individuals from 38 households were lost to follow up for participation in the contact survey, with a further three individuals declining consent.

### Demographic characteristics

The median age among household study participants was 15 years; 25% were younger than 7 years old and 75% were younger than 34 years old (Figure 1). There were notably fewer adult men than women. The male to female gender ratio among enrolled household members of all ages was 1:1.2, and 1:1.5 in adults. The respective gender ratio was 1:1.9 among the participants enrolled in the contact study and 1:1.9 among their reported contacts.

**Figure 1.**
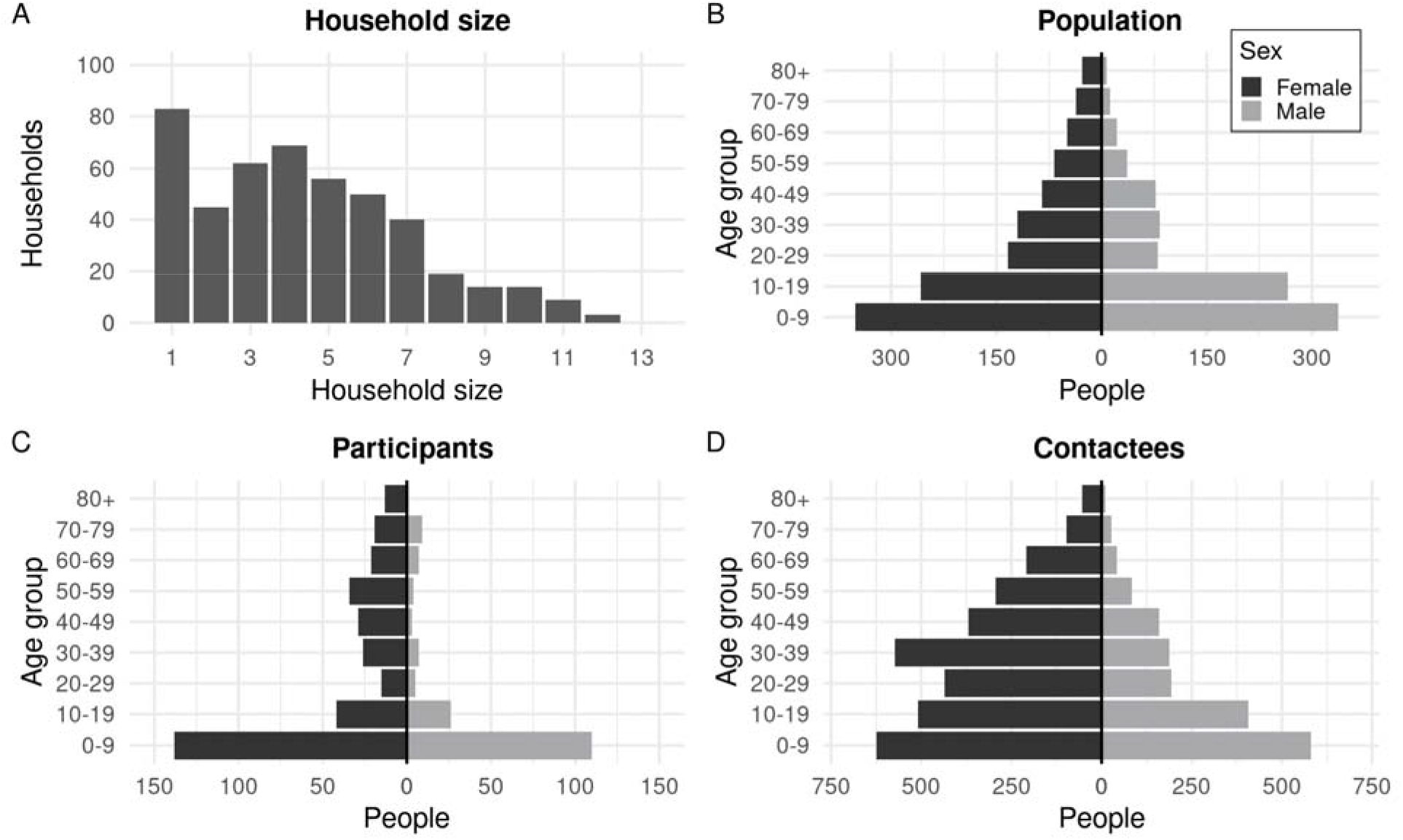
A. Frequency distribution of household sizes. B. age and sex distribution of all household members in participating households, which was used as the assumed population distribution. C. Age and sex distribution of participants included in the contact survey. D. Age and sex distribution of contactees listed by participants

The crude birth and death rates were estimated as 32 and 33 per 1,000 per year, respectively, in the six months preceding the survey. The crude under 5 years death rate was estimated at 57 per 1,000 per year. Crude in- and out-migration rates were high at 139 and 161 per 1,000 per year in the same period (Table 1). The majority of households (79%) settled in Digaale more than 3 years prior to the survey, while 6% settled in the year prior to the survey. The median household size was 4 individuals, ranging from 1 (83 households) to 12 (3 households) (Figure 1A).

**TABLE 1.** Characteristics of participating households and prevalence of risk factors in Digaale IDP camp

| Variable | N <sup>a</sup> | Estimate <sup>b</sup> |  |
| --- | --- | --- | --- |
| Demographic characteristics |  |  |  |
| Median household size | 464 | 4 | 2 - 6 (IQR) |
| Median age | 2049 | 15 | 7 - 34 (IQR) |
| Crude birth rate <sup>c</sup> | 33 | 31.9 | 26.4 - 38.7 |
| Crude death rate <sup>c</sup> | 34 | 32.9 | 27.2 - 39.7 |
| Crude U5 death rate <sup>c</sup> | 9 | 56.6 | 39.1 - 81.7 |
| Crude in-migration rate <sup>c</sup> | 144 | 139.3 | 127.1 - 152.7 |
| Crude out-migration rate <sup>c</sup> | 166 | 160.6 | 147.4 - 175 |
| Years since household settled in Digaale |  |  |  |
| <1 year | 27 | 5.8% | 4.6 - 7.1 |
| 1-2 years | 39 | 8.4% | 6.9 - 9.9 |
| 2-3 years | 31 | 6.7% | 5.3 - 8 |
| >3 years | 367 | 79.1% | 76.9 - 81.3 |
| Quality of shelters |  |  |  |
| Total number of rooms (mean) | 505 | 1.1 | 1.1 - 1.1 |
| Reported draught in shelter | 338 | 72.8% | 70.4 - 75.2 |
| Reported leakage in shelter | 341 | 73.5% | 71.1 - 75.9 |
| Indoor air pollution |  |  |  |
| Cooking fuel used |  |  |  |
| Charcoal | 275 | 59.3% | 56.8 - 61.9 |
| Firewood | 388 | 83.6% | 81.6 - 85.6 |
| Ventilation in cooking area |  |  |  |
| Cook outside | 308 | 66.4% | 63.8 - 68.9 |
| Ventilation absent | 107 | 23.1% | 20.8 - 25.3 |

| Variable | N<sup>a</sup> | Estimate<sup>b</sup> |  |
| --- | --- | --- | --- |
| Ventilation present | 49 | 10.6% | 8.9 - 12.2 |
| Primary water source |  |  |  |
| Tanker truck delivery | 456 | 98.3% | 97.6 - 99.0 |
| Rainwater collection | 7 | 1.5% | 0.9 - 2.2 |
| Substance use in household |  |  |  |
| None | 289 | 62.3% | 59.7 - 64.9 |
| Khat | 168 | 36.2% | 33.6 - 38.8 |
| Smoke | 126 | 27.2% | 24.8 - 29.6 |
| Snuff | 29 | 6.2% | 4.9 - 7.6 |
| Alcohol | 0 | 0% | 0 - 0 |
| Prevalence of malnutrition in U5 |  |  |  |
| Weight for age |  |  |  |
| Not underweight ($z > -2$) | 136 | 81.8% | 77.4 - 86.3 |
| Underweight ($z \leq -2$) | 27 | 14.1% | 10.1 - 18.1 |
| Severely underweight ($z \leq -3$) | 8 | 4.1% | 1.9 - 6.3 |
| Height for age |  |  |  |
| Not stunted ($z > -2$) | 124 | 71.7% | 65.8 - 77.6 |
| Stunted ($z \leq -2$) | 31 | 19.5% | 14.1 - 24.9 |
| Severely stunted ($z \leq -3$) | 16 | 8.8% | 5.3 - 12.2 |
| Weight for height |  |  |  |
| Not wasted ($z > -2$) | 150 | 89.2% | 85.5 - 92.9 |
| Wasted ($z \leq -2$) | 18 | 9.3% | 5.8 - 12.8 |
| Severely wasted ($z \leq -3$) | 3 | 1.5% | 0.1 - 2.8 |
| Middle-Upper Arm Circumference |  |  |  |
| Not wasted ($\geq 125\text{mm}$) | 165 | 96.8% | 94.5 - 99.1 |
| Wasted ($< 125\text{mm}$) | 5 | 3.2% | 0.9 - 5.5 |

| Variable | N<sup>a</sup> | Estimate<sup>b</sup> |  |
| --- | --- | --- | --- |
| Severely wasted (< 115mm) | 0 | 0% | 0 - 0 |
| Cumulative incidence of self-reported pneumonia |  |  |  |
| Ever |  |  |  |
| <2 years old | 39 | 54.3% | 45.7 - 62.9 |
| 2-5 years old | 62 | 51.4% | 43.4 - 59.5 |
| 6-14 years old | 25 | 29.1% | 19.2 - 39 |
| 15-29 years old | 18 | 33.4% | 20.8 - 46 |
| 30-49 years old | 15 | 27.2% | 14.7 - 39.6 |
| 50+ years old | 10 | 9.9% | 3.8 - 16.1 |
| In the last six months |  |  |  |
| <2 years old | 32 | 45.5% | 36.8 - 54.2 |
| 2-5 years old | 31 | 25.8% | 18.7 - 32.9 |
| 6-14 years old | 11 | 12.2% | 5.3 - 19.2 |
| 15-29 years old | 4 | 7.7% | 0.4 - 15 |
| 30-49 years old | 7 | 13.3% | 3.6 - 23.1 |
| 50+ years old | 3 | 2.8% | 0 - 6.4 |

<sup>a</sup> Total number of observations
<sup>b</sup> Central estimates are the weighted mean, median, or percentage. The uncertainty interval next to the central estimates is the 95% confidence interval for proportions and means, and interquartile range (IQR) for median estimates. A finite population correction was applied in calculating confidence intervals.
<sup>c</sup> Rates are per 1000 people per year

### Living conditions

The majority of households had only a single room (93%), with 6% having two rooms and 2% three or four rooms. Most households reported having draughts (73%) and leakage (74%) in their shelter. Both firewood (84%) and charcoal (60%) were commonly reported as cooking fuels. The majority of households usually cook outside (66%) or in a ventilated area inside (23%), with a minority (10%) of households reporting to cook in an unventilated area. Khat (36%) and tobacco (27%) were reported to be consumed by at least one household member in around one third of all households.

### Travel patterns

We estimate that just under half of the population never travels outside the Digaale camp, while one fifth does so at least once per week. Those who do travel predominantly go to nearby locations, including the city of Hargeisa. Less than 10% reported travelling further than 10km from the camp and less than 3% more than once a week (Table 2).

**TABLE 2.** Frequency of travel outside Digaale IDP camp

| **Travel distance** | **Most days of the week** |  | **At least once per week** |  | **At least once per month** |  | **Less than once per month** |  | **Never** |  |
| --- | --- | --- | --- | --- | --- | --- | --- | --- | --- | --- |
| < 5km | 17.7% | 13.6 - 21.8 | 21.4% | 17 - 25.8 | 12.7% | 9 - 16.2 | 4.0% | 2.2 - 5.9 | 44.2% | 39.2 - 49.2 |
| 5 - 10km | 8.3% | 5.3 - 12.0 | 11.4% | 8.1 - 14.7 | 16.7% | 13.1 - 20.4 | 6.2% | 3.7 - 8.7 | 57% | 51.8 - 62.3 |
| > 10km | 1.6% | 0 - 3.4 | 1.2% | 0 - 2.6 | 2.8% | 0.8 - 4.9 | 4.0% | 1.6 - 6.3 | 90.3% | 86.7 - 93.9 |

Estimates are the weighted proportion and corresponding 95% confidence interval. A finite population correction was applied in calculating confidence intervals.

### Anthropometric status

We estimate that 14% of all children under 5 years old in the population were underweight for their age, and 4% of all children severely underweight. Prevalence of global acute malnutrition (wasting, GAM) was 9% when expressed using weight-for-height Z score, with 2% severely malnourished. In contrast, GAM was 3% when assessed by MUAC, with no children severely malnourished and none diagnosed with oedema. 20% of children were stunted.

### Social contact patterns

The average number of direct daily contacts was relatively consistent by age (Table 3), and was highest for those aged 2-5 years (10.7) and lowest for those aged 50+ years (8.8). A large proportion (>77%) of these contacts are physical. The proportion of physical contacts is relatively consistent across contact frequency, duration, relationship, and setting (Figure 2). Nearly all direct contacts were made with previously known individuals (99% of all contacts), mostly household members (34% of all contacts), relatives not in the household (25%), and friends (30%). Most contacts are met daily or almost daily (88%), and most (42%) reported contacts lasted longer than four hours. All age groups report a high number of indirect casual contacts in addition to their direct contacts, with little variability across age-groups.

**TABLE 3.** Mean number of reported daily contacts by age, contact type and contact setting.

| **Age group** | **Contact type** |  |  | **Contact setting (Direct)<sup>a</sup>** |  |  |  |  |  |
| --- | --- | --- | --- | --- | --- | --- | --- | --- | --- |
|  | **Total (Direct)<sup>a</sup>** | **Physical (Direct)<sup>b</sup>** | **Total (Indirect)<sup>c</sup>** | **Home or another house** |  | **School or work** |  | **Other** |  |
| <2 | 9.3 8.9 - 9.8 | 6.7 6.1 - 7.3 | 11.5 10.5 - 12.5 | 8.7 | 8.3 - 9.2 | 0.1 | 0 - 0.1 | 0.6 | 0.4 - 0.8 |
| 2-5 | 10.7 10.2 - 11.2 | 8.6 8 - 9.2 | 11.9 11 - 12.8 | 10.1 | 9.6 - 10.6 | 0.2 | 0.1 - 0.3 | 0.6 | 0.4 - 0.8 |
| 6-14 | 10.3 9.3 - 11.3 | 8.4 7.4 - 9.5 | 13.4 12.1 - 14.6 | 7.4 | 6.5 - 8.3 | 2.3 | 1.5 - 3 | 0.8 | 0.5 - 1.2 |
| 15-29 | 8.9 8 - 9.9 | 7.1 6 - 8.3 | 14.0 12.3 - 15.6 | 6.0 | 5.1 - 6.9 | 1.2 | 0.6 - 1.8 | 2.1 | 1.3 - 2.8 |
| 30-49 | 10.3 9.1 - 11.5 | 7.6 6.4 - 8.9 | 14.6 12.6 - 16.5 | 7.4 | 6.3 - 8.5 | 1.2 | 0.1 - 2.2 | 1.8 | 0.9 - 2.7 |
| 50+ | 8.8 8.1 - 9.6 | 6.2 5.3 - 7 | 12.4 10.6 - 14.1 | 6.7 | 5.9 - 7.5 | 0.4 | 0.2 - 0.6 | 1.9 | 1.5 - 2.4 |

Estimates are the weighted mean and corresponding 95% confidence interval. A finite population correction was applied in calculating confidence intervals.
<sup>a</sup> Direct contacts were defined as in-person contacts with whom the participant had at least a short conversation.
<sup>b</sup> Physical contacts were direct contacts involving physical touch.
<sup>c</sup> Indirect contacts were additionally reported contacts that did not fit the definition of a direct contact.

**Figure 2.**
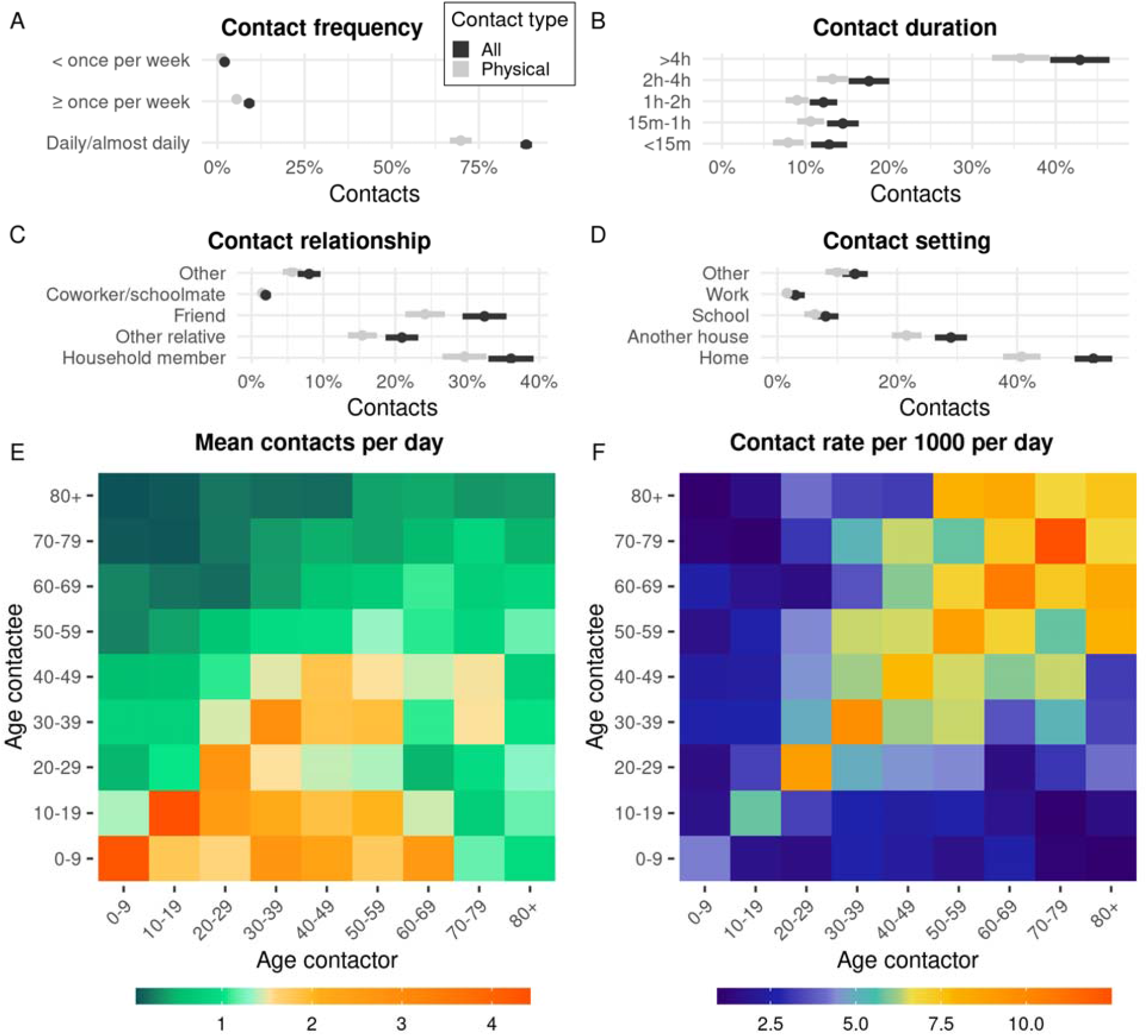
Panels show the reported frequency by which contactors meet contactees (A), the duration of contacts (B), the relationships between contactors and contactees (C), and the setting where contacts occurred (D). Direct contacts are stratified by contact type. Contact matrices show the weighted mean number of contacts per day contactors make with contactees of a certain age (E), and the age-specific weighted daily contact rates (reported per 1000 people; i.e. the rate at which any two individuals are assumed come into contact each day). Both matrices are adjusted for reciprocity of contacts.

The vast majority (>82%) of direct contacts are made at a home or in another house, with few reported contacts at school or work, or in other settings. The average number of school or work contacts is higher when restricted to participants who report at least one school or work contact, but remains lower compared to contacts made at home or in another house in all age groups (Supplemental table C1).

The contact matrices in Figure 2 show who contacts whom. We observe the highest average daily number of contacts within the same age group to be among children. The higher overall contact rates made by children reflect the relatively high proportion of the population that are children, while the per capita contact rates give an estimate of assortativity adjusted for population size. Overall, contacts were mostly age-assortative, especially in children, with more intergenerational contacts in adults. Out of 4,857 contacts, only 91 were reported to occur outside of Digaale, all of which occurred in Hargeisa. 95% uncertainty estimates around the contact rates are shown in Supplemental Figure C1.

Supplemental figure C2 shows contact patterns by setting. They are relatively age-assortative outside the household, while more age-disassortative within the household. Contacts were also gender-assortative (Supplemental figure C3). Children had higher contact rates with adult women than adult men, and age-assortative mixing was higher in men than in women.

### Self-reported pneumonia illness

There was a strong decreasing trend by age for self-reported cumulative incidence of pneumonia diagnosis (Table 1). Around 54% of children under 5 were reported to have ever been diagnosed with at least one episode of pneumonia, while 46% of children under 2 were reported to have been diagnosed with pneumonia within the last six months.

Using logistic regression, after adjusting for age and sex, we did not find evidence of any increase in the odds of cumulative incidence of self-reported pneumonia diagnosis for the majority of risk factors listed in Table 1 nor the total number of contacts (Supplement Section D). There was some evidence for an increased odds of self-reported pneumonia diagnosis in the six months preceding the survey for those living in households where snuff was consumed (OR 2.6; 95%CI 1.1 - 6.0).

## Discussion

To our best knowledge this is the first study to collect data on social contacts in any IDP camp, and to describe risk factors relevant for the spread of infectious respiratory diseases in a Somaliland IDP camp. We found that the majority of households have been living in Digaale for over three years, while estimated crude migration rates are relatively high. There is a high female to male ratio in adults living in Digaale.

We estimated a low mean age and corresponding high crude death rates, especially in those younger than five years. The estimated crude death rates (per 10,000 per day in the six months preceding the survey) was 0.9 for all ages and 1.5 for children younger than 5 years, and are considerably higher than those reported for Somalia including Somaliland between 2013-2018 (0.43 and 0.66).^25^ Our findings underscore the consistently higher mortality observed among IDPs, compared to other population groups.^26^ Although we do not know the aetiology of these deaths, a high proportion of participants reported a historical pneumonia diagnosis, especially in younger children.

Several known risk factors for respiratory infections in children are prevalent.^27,28^ First, approximately 14% of children in Digaale were underweight. Although no children were found to have severe acute malnutrition, food insecurity may vary substantially over time. Nearly a fifth of children had stunted growth, reflecting long-term undernutrition, which may indicate periods of inadequate access to food for their families. These data are similar to estimates in IDP populations in Somalia.^29^

Second, individuals live in relatively poor-quality shelters, while local minimum temperatures can drop to 5 °C. Firewood and charcoal are the only cooking fuels used in Digaale, and both can raise levels of indoor air pollution. However, only a small proportion of households cooks in an unventilated area indoors, which mitigates this risk. At least one household member smokes in one third of all households, which could further affect levels of indoor air pollution.

Third, levels of crowding within Digaale are substantial. While the average household size of four people is below the national average^30^, most households share only a single small room. Increased human contact facilitates the transmission of pathogens causing respiratory diseases. On average, individuals have around 10 direct contacts each day, which is consistent between age groups, and the majority of these contacts involve physical touch. The average number of reported direct contacts was lower in this setting when compared to contact surveys conducted in Kenya^7^ or a South African township^9^, though this likely reflect a difference in survey design, as we excluded casual contacts from our contact definition. In Digaale, individuals reported on average a further 13 indirect contacts per day, though these are less likely to result in respiratory transmission.

The average proportion of direct contacts that were physical was higher in Digaale (78%) compared to a crowded township in South Africa (27%)^9^, and a peri-urban township in Zimbabwe (57%)^8^, but similar to a rural setting in Uganda (73%).^6^ Compared to these settings, we find that contacts in Digaale are more likely to occur at the home, with very few made at school or work.

There are several limitations to our study. First, our estimates may be affected by selection bias, as the proportion of male participants of working age included in the survey was smaller than the proportion of men of this age in the population. We could only conduct data collection during daylight hours, and may therefore have missed individuals who work outside Digaale, as many leave the camp very early in the morning and only return late at night. Of the 35% of shelters not included in the survey, many were (according to neighbours) single occupancy dwellings whose residents were absent during working hours.

Second, caregivers completed the survey on behalf of young children, which may have resulted in underreporting of non-household contacts of those children, and explain why e.g. reported direct contacts at school are very low.

Third, we designed the survey to collect retrospective data on social contacts, which could result in a lower reported number of contacts due to recall bias^10^. We asked participants to remember their contacts one day in advance, and used structured interviews with specific prompts in an attempt to minimize any recall issues. We assessed contact rates for potential underreporting by comparing the reported intra-household contact rates with the expected intra-household contact rates based on household demographics (Supplemental Figure C4). While the relative reported contact patterns were very similar to the expected contact patterns, their absolute values were lower, with the dominant eigenvalue of the matrix 35% lower than that of the expected matrix. This may reflect underreporting of household contacts, but could also reflect household members having less than one contact per day, on average.

Finally, while our regression analysis did not find any significant association between any of the known risk factors considered and self-reported estimates of pneumonia, our study was not designed nor powered to do so. Self-reported pneumonia may be an unreliable outcome measure, and it was unclear what case definitions were used when making the diagnoses.

## Conclusion

We find a high prevalence of risk factors for lower respiratory tract infections in an IDP camp in Somaliland. Crude death rates in the camp exceeded the already high rates in the host population Somalia and Somaliland. Compared to other settings, a large degree of contacts are of a physical nature, and the vast majority of contacts are made within homes. We find social mixing to be assortative by both age and sex, but there is low variability in total number of contacts by age. Malnutrition is prevalent, and indoor air pollution is likely high, while individuals live in crowded shelters of poor quality. This study illustrates that such IDP settings potentially present a perfect storm of risk factors for lower respiratory tract infections and their transmission, often combined with inadequate access to curative or preventive health care. Innovative approaches to address such risks are urgently needed.

## Data Availability

The computer code and anonymized data used to conduct analyses for this article are available on GitHub.

https://github.com/kevinvzandvoort/espicc-somaliland-digaale-survey-2019

## Acknowledgements

The authors thank Hamse Shaban Ahmed, Ayaan Ismail Adan, Khadar Abiib Ahmed, Ayaan Mohamoud Ali, Hamda Awil Garaad, Ahmed Yusuf Mohamed, Suaad Abdi Osman, Amiin Abdi Ismail, Nimco Abdilahi Ismail, AbdiFatah Mohamed Ahmed, Mustafe Shugri Mohamed, and Nimco Mohamed Abdi from the Republic of Somaliland Ministry of Health Development for their data collection efforts. Electronic data solutions were provided by LSHTM Open Data Kit (odk.lshtm.ac.uk).

