## Supplementary Material for "Social contacts and other risk factors for respiratory infections among internally displaced people in Somaliland"

### Supplemental Material

This supplemental material includes four sections:

A. Photos of Digaale IDP camp
B. Assessment of missing households and FPC
C. Additional analyses of contact patterns
D. Logistic regression analysis

Additional analysis scripts, anonymized data, and questionnaire scripts are available on GitHub via <https://github.com/kevinvzandvoort/espicc_somaliland_digaale_contact_survey_2019>.

#### Section A. Photos of Digaale IDP camp

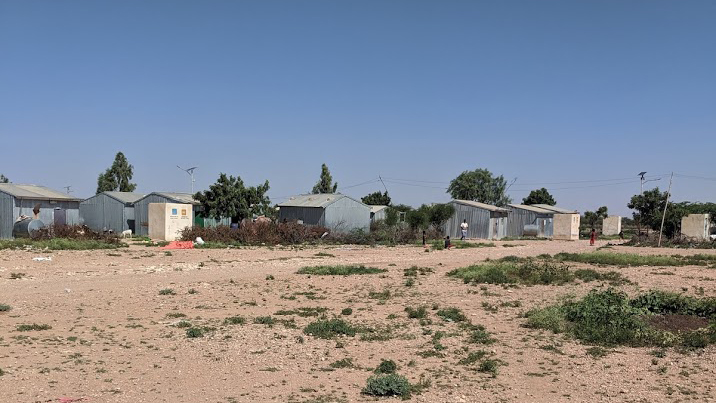

Several shelters seen from a distance near the border of Digaale.

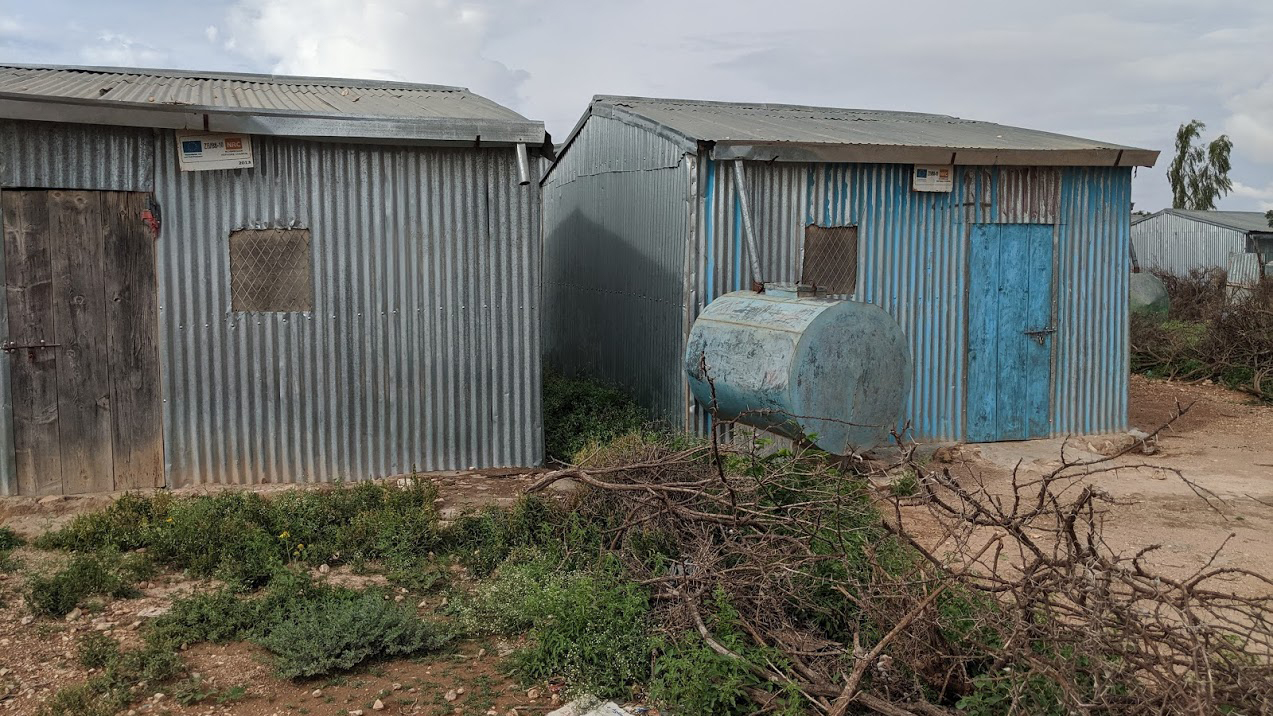

Two shelters seen from the front. All shelters are made from corrugated sheets. Several shelters have not been inhabited for some time.

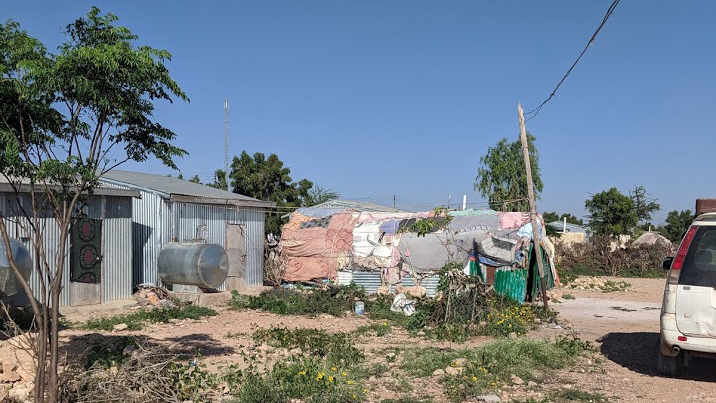

Two shelters in Digaale. There is a water tank on the left-hand side of each shelter. A traditional Somali hut has been constructed next to the right shelter to increase space.

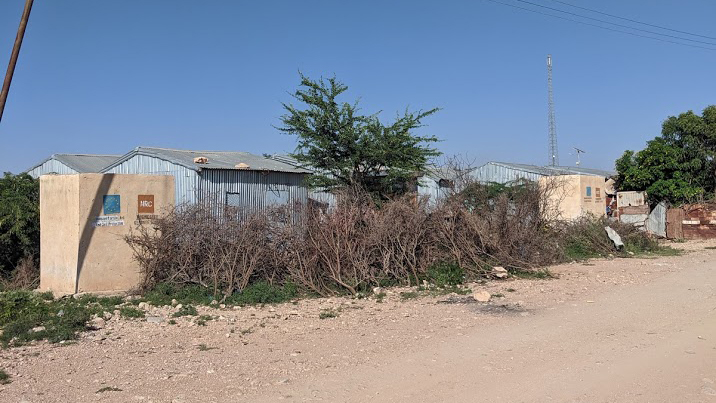

The yellow concrete blocks are latrines. Each shelter has its own private latrine.

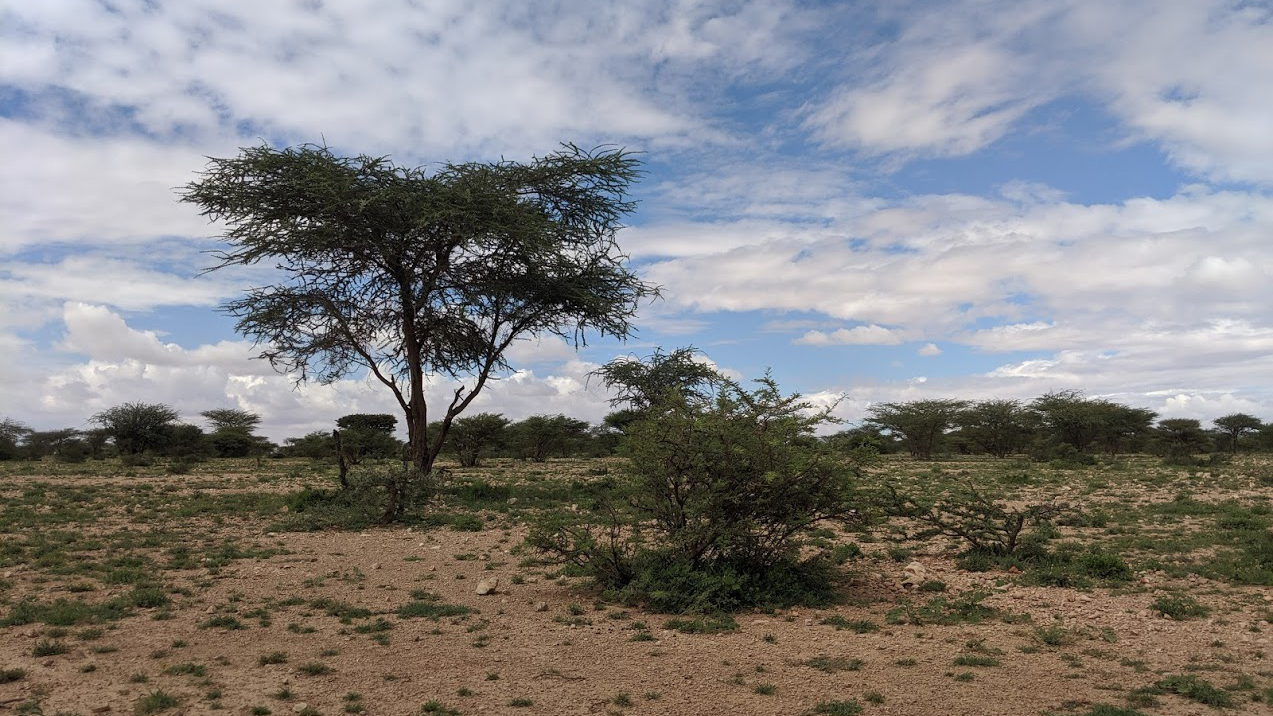

Digaale is surrounded by desert and shrubland.

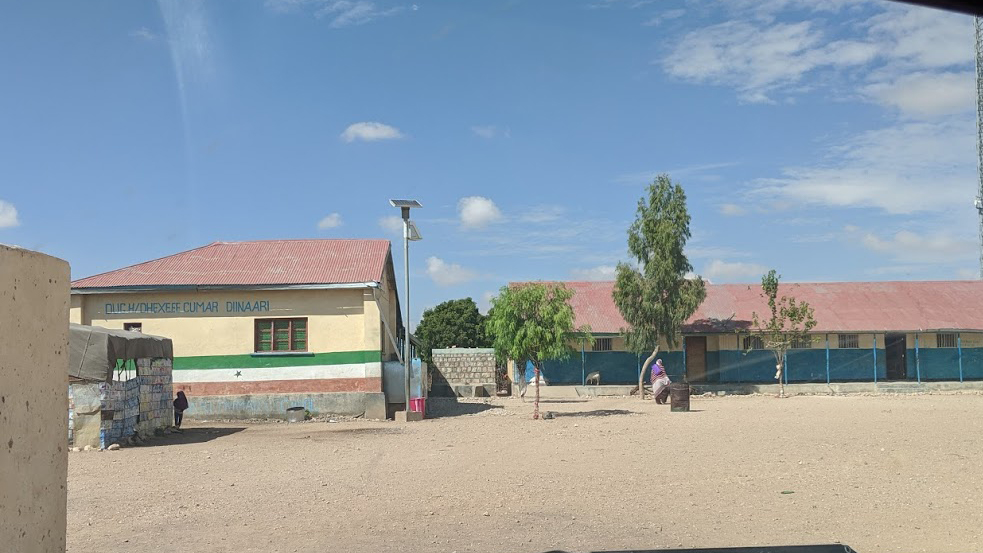

There is a primary school in the center of Digaale.

#### Section B. Assessment of missing households and FPC

Supplemental Table B1 shows the results of an assessment of a random sample of 96 shelters of a total of 405 shelters where no individual was present on multiple visits. We asked people living in neighbouring shelters whether they knew if the shelter was occupied. We could not retrieve any information for 23 shelters. Of the 73 shelters for which we did retrieve information, 36 were listed as having been vacant for a long time (empty). Twenty-eight were occupied, but residents either only returned back to Digaale at night, or were travelling to care for animals. Nine were no unique shelters, but were either a shop or the shelter was combined with a different shelter already included in the survey.

Using these data, we can estimate population totals and use the lower bound of these estimates to conservatively assume that at least 157 shelters are empty and at least 22 shelters are a shop or existing household. Thus we assume that out of all 894 shelters in Digaale, only 715 are inhabited with unique households (Supplemental Table B2).

| **SUPPLEMENTAL TABLE B1.** Reported status of 73 shelters from a random sample of shelters where no individual was present on multiple visits | | | | | |
| --- | --- | --- | --- | --- | --- |
| **Status of shelters not present** ^a^ | **n** ^b^ | **%** ^c^ | **(95% CI)** | **N** ^d^ | **(95% CI)** |
| Empty | 36 | 49.3% | 38.9 - 59.8 | 200 | 157 - 242 |
| Occupied | 28 | 38.4% | 28.2 - 48.5 | 155 | 114 - 197 |
| Shop or same household | 9 | 12.3% | 5.5 - 19.2 | 50 | 22 - 78 |
| Total | 73 |  |  | 405 |  |
| ^a^ As reported by individuals living in neighbouring shelters.  ^b^ Number of recorded observations.  ^c^ Estimated proportion of shelters in Digaale with this status.  ^d^ Estimated total number of shelters in Digaale with this status. | | | | | |

Of the 715 households that we assumed to be inhabited (Supplemental Table B2), we visited 489 who were all invited to participate in the study. Most (464) of these households consented to be part of the survey. Assuming that the demographics in the households that were included in the survey were similar to those of the inhabited households not included in the survey, we calculated a correction factor for the population size as 715/464 which was used when calculating the finite population corrections in our analyses.

| **SUPPLEMENTAL TABLE B2.** Total number of shelters and households in Digaale and included in the survey | | |
| --- | --- | --- |
| **Shelters** | **N** | **%** |
| Total shelters in Digaale | 894 |  |
| Assumed inhabited shelters in Digaale | 715 | 80% ^a^ |
| Household present during survey | 489 | 68% ^b^ |
| Households consented that were present | 464 | 95% ^c^ |
| ^a^ Percentage of total shelters that is assumed to be inhabited.  ^b^ Percentage of total assumed inhabited shelters where a household was present and visited during the survey.  ^c^ Percentage of present households that consented to participate. | | |

#### Section C. Additional analyses of contact patterns

To assess uncertainty in the contact matrix, we took 10,000 bootstrap samples where we resampled participants in the contact data with replacement. We recalculated sample weights in each bootstrap sample, and calculated the corresponding contact matrix. We then calculated the dominant eigenvalue for each bootstrapped contact matrix, and selected the contact matrices whose dominant eigenvalue lay closest to the mean, 2.5th percentile, and 97.5th percentile of all eigenvalues. These matrices are represented as the mean, lower, and higher contact matrix in Supplemental Figure C1.

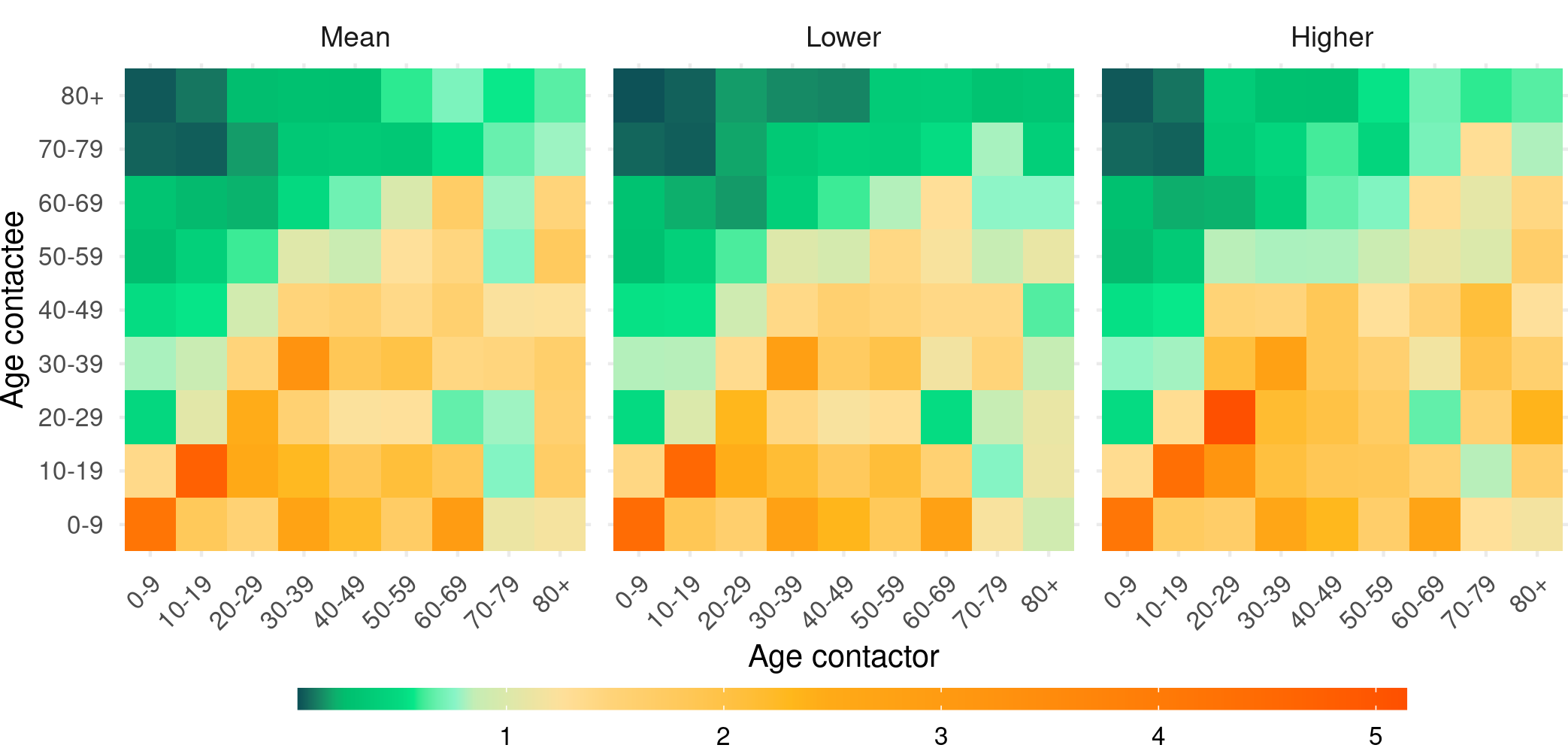

**SUPPLEMENTAL FIGURE C1.**	Mean daily age-specific contact matrix and lower and upper 95% uncertainty estimates of age dependent social contact matrices adjusted for reciprocity of contacts in Digaale IDP camp, Somaliland

Supplemental Figure C2 shows the age-specific contact matrix and per-capita age specific contact matrix for contacts made with (intra-household) and without (extra-household) household members. While extra-household contacts are very assortative, intra-household contacts are of more homogeneous nature.

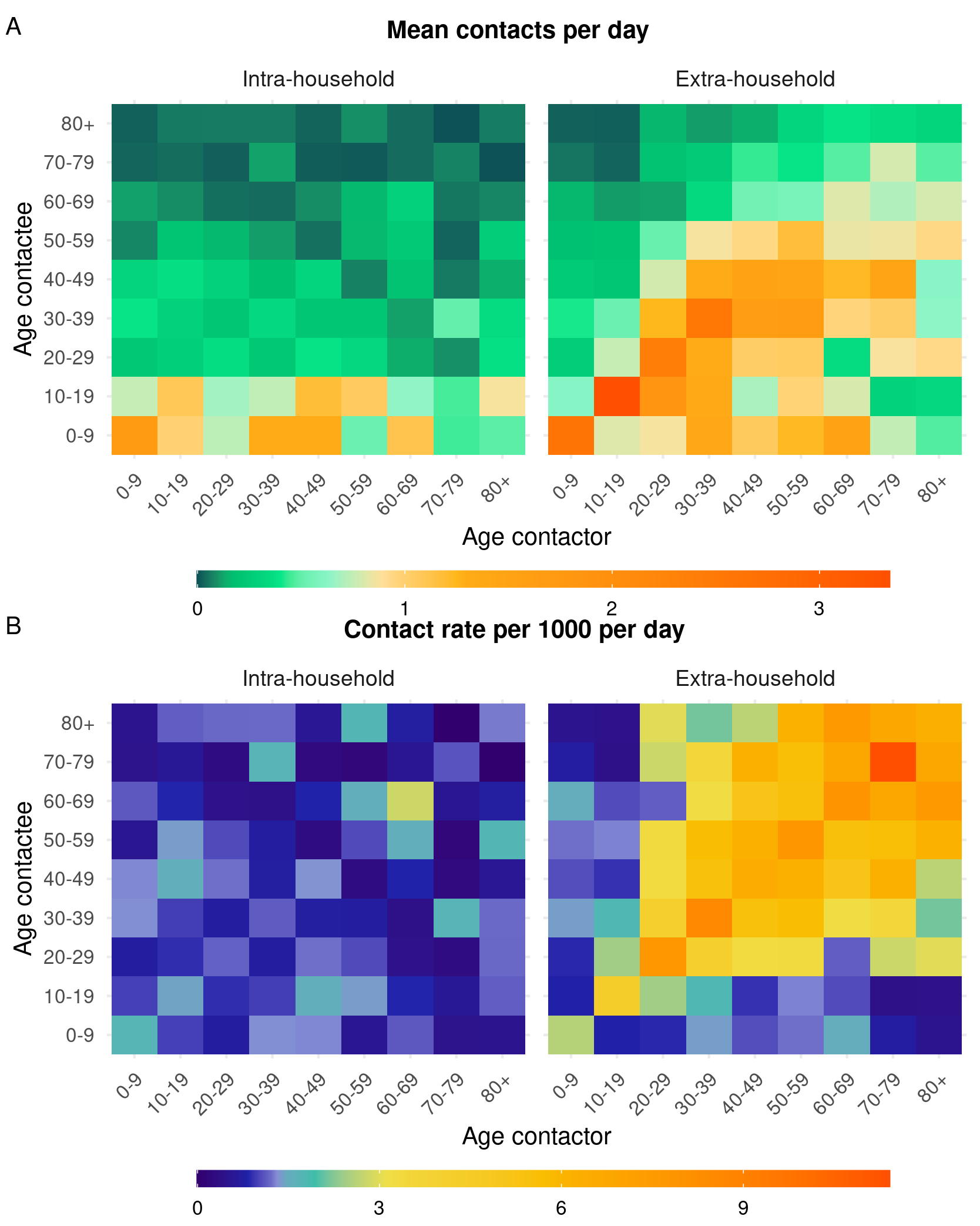

**SUPPLEMENTAL FIGURE C2.**	Weighted mean number of contacts per day between contactors and contactees of certain ages (A), and age-specific weighted daily contact rates (reported per 1000 people) for contacts with household members (intra-household) and non-household members (extra-household) in Digaale IDP camp, Somaliland. Both matrices are adjusted for reciprocity of contacts

Supplemental Figure C3 shows daily contact rates by age and gender. Contact rates are assortative by both age and gender, with less mixing between the two genders compared to within each gender. Female adults make more contacts with children compared to male adults.

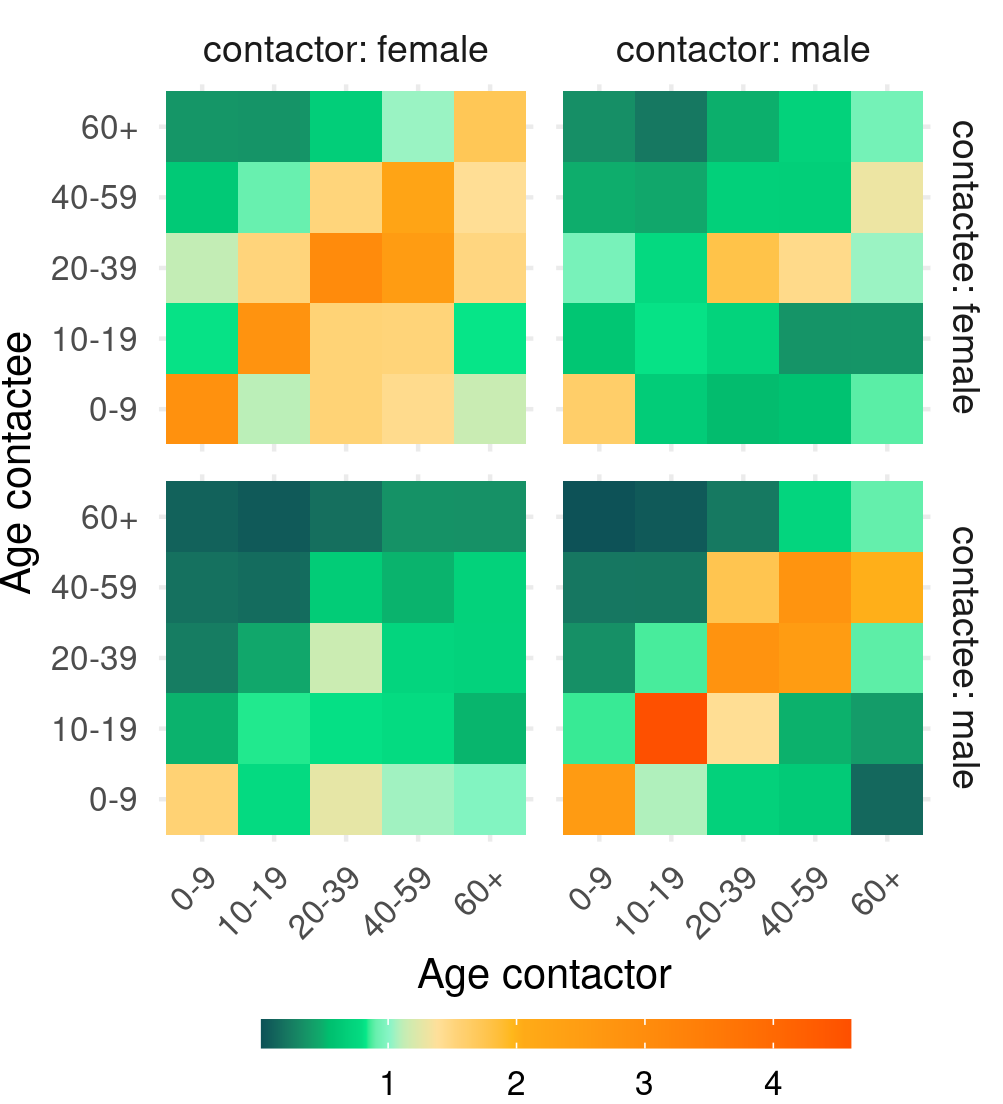
 **SUPPLEMENTAL FIGURE C3.**	Daily contact rates ^a^ by age and gender
^a^ Matrices are adjusted for reciprocity, meaning that e.g. the total number of contacts of females aged j with males aged i are the same as the total number of contacts of males aged i with females aged j.

Supplemental Figure C4 shows the contacts made with household members as reported by participants in the survey (reported), and as expected by assuming that all individuals who live in the same household would all make a single contact per day. Overall patterns are very similar, but contact rates are generally lower in the empirical data compared to the expected data. The dominant eigenvalue of the matrix based on the empirical data is 35% lower than that of the matrix with expected intra-household contacts.

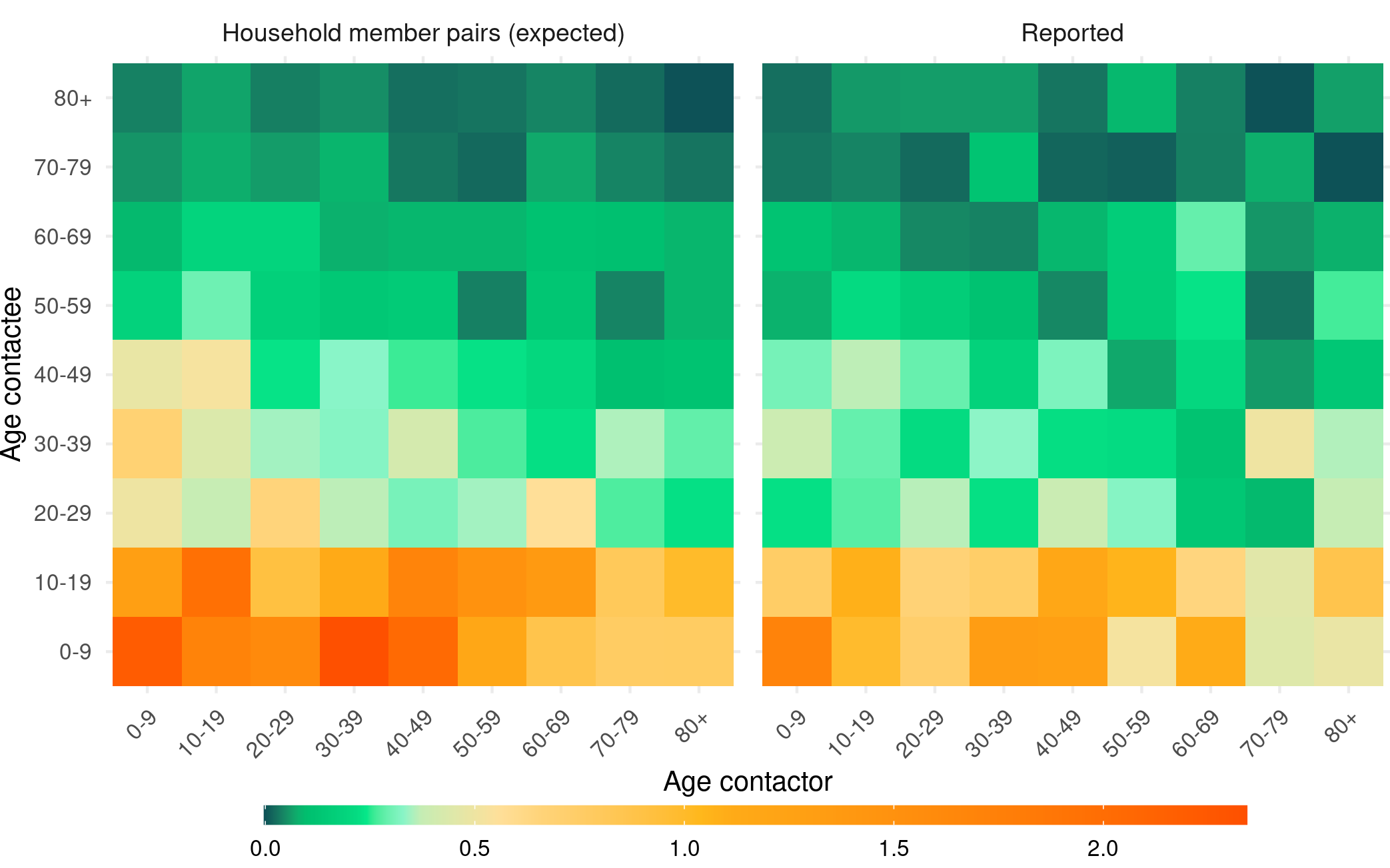

**SUPPLEMENTAL FIGURE C4.**	Expected ^a^ and observed contact matrices for contacts made with household members
^a^ Expected values are reported for all contact members for which data is available, and calculated by assuming that all individuals living in the same household would report one contact per day.

Supplemental Table C1 shows the estimated average daily contact rate at school or work by age, excluding individuals who reported no work or school contacts. Estimates are substantially higher in all age group compared to estimates where contactors with 0 contacts are not excluded (Table 3 in main manuscript).

| **SUPPLEMENTAL TABLE C1.** Average daily number of (direct) contacts reported at school or work settings, excluding individuals who reported no school and work contacts | | |
| --- | --- | --- |
| **Age group** | **Total (Direct) contacts at school or work** | |
| <2 | 1.0 | 1 - 1 |
| 2-5 | 3.4 | 2.5 - 4.2 |
| 6-14 | 5.1 | 4.1 - 6.2 |
| 15-29 | 3.4 | 2.3 - 4.6 |
| 30-49 | 5.8 | 2.4 - 9.2 |
| 50+ | 1.9 | 1.1 - 2.7 |
| Estimates are the weighted mean and 95% confidence interval. | | |

#### Section D. Logistic regression analysis

We assessed the relationship between several household- and individual level risk factors with self-reported cumulative pneumonia incidence in the six months preceding the survey (Supplemental Table D1), or ever (Supplemental Table D2) through logistic regression. We decided to adjust all risk factors for age and sex a priori.

The cumulative incidence patterns show a strong non-linear trend by age (Table 1 in main manuscript). A likelihood ratio test found good evidence to include age as a categorical variable (p = 0.012) for models regressing risk factors against cumulative pneumonia incidence in the last six months (Supplemental Table D1), but not for models regressing against cumulative pneumonia incidence ever (Supplemental Table D2). Age group was included as a categorical variable for both outcomes. In models where data was restricted to children younger than five years of age (those that include anthropometric estimates), age in months was used as a continuous variable instead.

| **SUPPLEMENTAL TABLE D1.** Association between risk factors and self-reported pneumonia diagnosis in the six months preceding the survey | | | | |
| --- | --- | --- | --- | --- |
| **Variable** | | **OR** ^a^ | **95% CI** | **p-value** |
| Demographic characteristics | | | | |
| Household size | | 1.06 | 0.95 - 1.18 | 0.274 |
| Household members <5y | | 1.05 | 0.78 - 1.4 | 0.764 |
| Household members <2y | | 0.81 | 0.46 - 1.4 | 0.469 |
| Years since household settled in Digaale | | | | |
| >3 years | | *ref* |  |  |
| 2-3 years | | 0.63 | 0.22 - 1.59 | 0.361 |
| 1-2 years | | 0.71 | 0.3 - 1.57 | 0.419 |
| <1 year | | 0.82 | 0.3 - 2.02 | 0.674 |
| Quality of shelter | | | | |
| Total number of rooms | | 1.03 | 0.49 - 1.91 | 0.929 |
| Reported draught in shelter | |  |  |  |
|  | no | *ref* |  |  |
|  | yes | 0.81 | 0.48 - 1.38 | 0.421 |
| Reported leakage in shelter | |  |  |  |
|  | no | *ref* |  |  |
|  | yes | 1.35 | 0.76 - 2.5 | 0.322 |
| Indoor air pollution | | | | |
| Use charcoal for cooking | |  |  |  |
|  | no | *ref* |  |  |
|  | yes | 0.93 | 0.56 - 1.58 | 0.798 |
| Use firewood for cooking | |  |  |  |
|  | no | *ref* |  |  |
|  | yes | 1.05 | 0.56 - 2.04 | 0.889 |
| Use ventilation when cooking | |  |  |  |
|  | no | *ref* |  |  |
|  | yes | 2.77 | 0.86 - 12.43 | 0.121 |
|  | cook outside | 3.63 | 1.23 - 15.6 | 0.039 |
| Substance use in household | | | | |
| Household member who uses khat | |  |  |  |
|  | no | *ref* |  |  |
|  | yes | 1.07 | 0.64 - 1.79 | 0.781 |
| Household member who smokes | |  |  |  |
|  | no | *ref* |  |  |
|  | yes | 0.99 | 0.57 - 1.7 | 0.978 |
| Household member who uses snuff | |  |  |  |
|  | no | *ref* |  |  |
|  | yes | 2.64 | 1.12 - 6.01 | 0.023 |
| Contact behaviour | | | | |
| Total number of direct contacts | | 0.99 | 0.92 - 1.07 | 0.785 |
| Total number of physical contacts | | 1.02 | 0.96 - 1.1 | 0.503 |
| Total number of contacts at home | | 1.00 | 0.92 - 1.07 | 0.919 |
| Total number of contacts at school | | 0.82 | 0.59 - 1.02 | 0.138 |
| Total number of contacts at work | | 0.62 | 0.17 - 1.1 | 0.307 |
| Total number of contacts at other settings | | 1.06 | 0.93 - 1.2 | 0.355 |
| Malnutrition in U5 | | | | |
| Weight by age | |  |  |  |
|  | Not underweight (z > -2) | *ref* |  |  |
|  | Underweight (z ≤ -2) | 1.12 | 0.41 - 2.92 | 0.818 |
|  | Underweight (z ≤ -3) | 0.35 | 0.02 - 2.33 | 0.347 |
| Height by age | |  |  |  |
|  | Not stunted (z > -2) | *ref* |  |  |
|  | Stunted (z ≤ -2) | 0.76 | 0.29 - 1.89 | 0.573 |
|  | Severely stunted (z ≤ -3) | 1.20 | 0.36 - 3.81 | 0.755 |
| Weight by height | |  |  |  |
|  | Not wasted (z > -2) | *ref* |  |  |
|  | Wasted (z ≤ -2) | 0.47 | 0.1 - 1.65 | 0.278 |
|  | Severely wasted (z ≤ -3) | 0.00 |  |  |
| Middle-Upper Arm Circumference | |  |  |  |
|  | Not wasted (≥ 125mm) | *ref* |  |  |
|  | Wasted (< -125mm) | 1.84 | 0.28 - 14.85 | 0.521 |
| ^a^ All odds ratios are adjusted for age and sex. | | | | |

| **SUPPLEMENTAL TABLE D2.** Association between risk factors and self-reported pneumonia diagnosis (ever) | | | | |
| --- | --- | --- | --- | --- |
| **Variable** | | **OR** ^a^ | **95% CI** | **p-value** |
| Demographic characteristics | | | | |
| Household size | | 1.03 | 0.94 - 1.12 | 0.567 |
| Household members <5y | | 0.98 | 0.76 - 1.25 | 0.845 |
| Household members <2y | | 0.65 | 0.4 - 1.04 | 0.079 |
| Years since household settled in Digaale | | | | |
| >3 years | | *ref* |  |  |
| 2-3 years | | 0.41 | 0.17 - 0.9 | 0.034 |
| 1-2 years | | 0.57 | 0.28 - 1.12 | 0.111 |
| <1 year | | 0.71 | 0.31 - 1.58 | 0.413 |
| Quality of shelter | | | | |
| Total number of rooms | | 0.81 | 0.44 - 1.4 | 0.467 |
| Reported draught in shelter | |  |  |  |
|  | no | *ref* |  |  |
|  | yes | 0.87 | 0.56 - 1.35 | 0.526 |
| Reported leakage in shelter | |  |  |  |
|  | no | *ref* |  |  |
|  | yes | 0.98 | 0.62 - 1.56 | 0.934 |
| Indoor air pollution | | | | |
| Use charcoal for cooking | |  |  |  |
|  | no | *ref* |  |  |
|  | yes | 0.86 | 0.56 - 1.31 | 0.479 |
| Use firewood for cooking | |  |  |  |
|  | no | *ref* |  |  |
|  | yes | 1.04 | 0.62 - 1.75 | 0.886 |
| Use ventilation when cooking | |  |  |  |
|  | no | *ref* |  |  |
|  | yes | 0.86 | 0.42 - 1.82 | 0.693 |
|  | cook outside | 1.04 | 0.54 - 2.04 | 0.910 |
| Substance use in household | | | | |
| Household member who uses khat | |  |  |  |
|  | no | *ref* |  |  |
|  | yes | 0.83 | 0.54 - 1.26 | 0.388 |
| Household member who smokes | |  |  |  |
|  | no | *ref* |  |  |
|  | yes | 0.61 | 0.38 - 0.96 | 0.035 |
| Household member who uses snuff | |  |  |  |
|  | no | *ref* |  |  |
|  | yes | 2.12 | 0.97 - 4.69 | 0.060 |
| Contact behaviour | | | | |
| Total number of direct contacts | | 1.01 | 0.95 - 1.07 | 0.714 |
| Total number of physical contacts | | 1.02 | 0.97 - 1.08 | 0.412 |
| Total number of contacts at home | | 1.02 | 0.96 - 1.08 | 0.568 |
| Total number of contacts at school | | 0.98 | 0.86 - 1.11 | 0.779 |
| Total number of contacts at work | | 1.02 | 0.8 - 1.25 | 0.858 |
| Total number of contacts at other settings | | 0.99 | 0.89 - 1.1 | 0.904 |
| Malnutrition in U5 | | | | |
| Weight by age | |  |  |  |
|  | Not underweight (z > -2) | *ref* |  |  |
|  | Underweight (z ≤ -2) | 1.66 | 0.65 - 4.54 | 0.302 |
|  | Underweight (z ≤ -3) | 0.44 | 0.06 - 2.37 | 0.359 |
| Height by age | |  |  |  |
|  | Not stunted (z > -2) | *ref* |  |  |
|  | Stunted (z ≤ -2) | 1.18 | 0.51 - 2.81 | 0.697 |
|  | Severely stunted (z ≤ -3) | 1.26 | 0.4 - 4.15 | 0.688 |
| Weight by height | |  |  |  |
|  | Not wasted (z > -2) | *ref* |  |  |
|  | Wasted (z ≤ -2) | 0.80 | 0.26 - 2.47 | 0.692 |
|  | Severely wasted (z ≤ -3) | 0.00 |  |  |
| Middle-Upper Arm Circumference | |  |  |  |
|  | Not wasted (≥ 125mm) | *ref* |  |  |
|  | Wasted (< -125mm) | 1.53 | 0.24 - 12.26 | 0.657 |
| ^a^ All odds ratios are adjusted for age and sex. | | | | |
